# Impaired Proactive Cognitive Control in Parkinson’s disease

**DOI:** 10.1101/2023.04.14.23288567

**Authors:** Julius Kricheldorff, Julia Ficke, Stefan Debener, Karsten Witt

## Abstract

Adaptive control has been studied in Parkinson’s disease (PD) mainly in the context of proactive control and with mixed results. We compared reactive- and proactive control in 30 participants with Parkinson’s disease (PD) to 30 age matched healthy control participants (HC). The electroencephalographic (EEG) activity of the participants was recorded over 128 channels while they performed a numerical Stroop task, in which we controlled for confounding stimulus-response learning. We assessed effects of reactive- and proactive control on reaction time-, accuracy- and EEG time-frequency data. Behavioral results show distinct impairments of proactive-reactive control in participants with PD, when tested on their usual medication. Participants with PD were unable to adapt cognitive control proactively and were less effective to resolve conflict using reactive control. Successful reactive and proactive control in the HC group was accompanied by a reduced conflict effect between congruent and incongruent items in midline-frontal theta power. Our findings provide evidence for a general impairment of proactive control in PD and suggest that the same may be the case for reactive control.

## Introduction

Parkinson’s disease (PD) is a neurodegenerative disorder primarily diagnosed and characterized by symptoms causing impaired motor functioning. PD also negatively affects multiple domains of cognition. Cognitive deficits associated with PD have been demonstrated to be an even larger detriment to quality of life of patients with PD’ than motor impairments(1). Individuals with PD display deficits in inhibition (e.g. 2,3), reinforcement learning (4), and cognitive control (5). Further, there is conflicting evidence whether individuals with PD exhibit a diminished adaptive control capacity (6). Adaptive control is the ability to adjust cognitive control to a given context and is thought to be essential for successful goal direct behavior in dynamic environments (7,8). Therefore, gaining a better understanding of how PD affects adaptive control is crucial.

Cognitive control describes the ability to regulate thoughts, or behavior to align with internal behavioral goals (9) and is often measured using interference tasks. For example, in the classical Stroop task (10) participants have to name the color that a particular word is printed in. This is relatively easy if the color and the word agree (congruent/no-conflict) - for example, the word RED, printed in red. The task becomes more difficult, when the color and word oppose each other (conflict/incongruent trial) - for example the word GREEN printed in the color red. Here the automatic tendency of the learned reading behavior, may interfere with the task of naming the color. Cognitive control is required to resolve the ensuing conflict/interference. Cognitive control can be measured by the strength of the interference effect. The interference effect measures the difference on reaction time or error rate between trials that contain conflicting/incongruent information versus trials containing non-conflict/congruent information. The dual mechanisms of control (DMC) framework (8,9) posits two modes of cognitive control, proactive- and reactive control. Proactive control is effortful, sustained over time and is thought to be already active before conflict is encountered. Reactive control is a “late-correction” mechanism (9). It is only engaged after encountering conflict. Proactive control is considered costly and resource intensive, making reactive control the default option (9).

### Measuring Adaptive Control

Adaptive control (also control learning or context-control learning) describes the regulation/adaptation of cognitive control resources to stable contexts repeatedly experienced over time. A common method to measure proactive control adaptation is to compare the strength of the interference effect in lists containing primarily conflicting (MI) items, in contrasts to list containing primarily non-conflicting (MC) items (11) - termed “list-wide proportion congruency effect” (LWPCE). The frequent exposure to conflict, in the MI list containing high proportion of conflicting items, should lead to the implementation of a “global control” adaptation (11). Cognitive control is sustained over the whole list/block (i.e. a temporal context) in anticipation of upcoming conflicts. Thus, participants are able to generalize the learned temporal context to other instances or items (12). This is reflected in a reduction in interference. Control shifts attention toward the response predicting stimulus feature (e.g. the color word RED), away from the conflicting feature (e.g. the color of the word green). This allows participants to respond relatively faster and more accurately in trials containing conflicting information. Moreover, typically reaction times (RTs) slow down in non-conflict trials. Since attention is shifted away from the conflicting stimulus feature (e.g. the color) it cannot facilitate the response. Thus, instead of two predictive stimulus features participants base their responses largely on one stimulus feature in the non-conflict case. In the context of MC lists control is seldom required and only applied reactively. Less control leads to more interference by the conflicting stimulus feature (slowing in RT and increase in error rates) on conflict/incongruent trials. Due to facilitation by both stimulus features RTs and error rates decrease on congruent trials. The difference in interference effects in high- (MI) versus low (MC) conflict context can be used to quantify the proactive control adaptation.

Reactive control adaptations can be measured by manipulating conflict proportions at the item level - termed “item-specific proportion congruency effect” (ISPCE) (11). Similar to the LWPCE, specific items are presented more often containing conflicting information (MI), and others are presented more often not containing conflicting information (MC). In contrast to the LWPCE, the temporal context (e.g. lists/blocks) is balanced in terms of conflict presentation. Hence, the manipulation occurs only “locally” on the level of the items. Due to the balanced overall presentation of conflict trials, participants are unable to predict conflict in the next item. Hence, conflict adaptation can only occur bottom-up (12) or reactively after encountering the item with its conflict identifying predictive feature. Thus, participants are not learning a “global” temporal context-association but a “local” feature-association, specific to a set of items. Reactive control is then similarly estimated by calculating the difference in interference between MC-Items and MI-Items.

Measurements of cognitive control adaptations are often criticized as the experimentally induced effects can also be explained by simpler stimulus-response (S-R) learning without the need to invoke adaptation of cognitive control mechanisms (for a review see 13). To isolate adaptive control processes independently of S-R learning, Braem et al. (11) recommend to induce adaptive control processes in one set of items, and measure the effects in a second set of unbiased items. Here we refer the former as inducer items and the latter diagnostic items (for an overview of how to employ these manipulations in different experimental contexts, see 11).

### Adaptive Control and Parkinson’s Disease

The literature on adaptive control abilities in PD is ambiguous, showing both evidence in favor of intact- and impaired adaptive control. Proactive control adaptation in PD has often been assessed using congruency sequence effect (CSE) manipulations. The CSE can be considered a more local, transient measure of proactive control (11) where adaptation in response to conflict in the previous trial is evaluated (14). Participants with PD on their usual dopaminergic medication (DOPA ON), in contrast to healthy control (HC) participants, have been reported to show comparable CSE modulations on RT (15) and no CSE modulations (16,17) on RT, saccadic latencies, and N2 - and lateralized readiness potential. Global proactive control adaptation, as measured by the LWPCE has been reported to be absent in PD in one study (6). Successful adaptation of proactive control may also depend on dopamine replacement therapy. Duthoo et al. (18) found the CSE to be impaired in participants with PD ON-their dopaminergic medication, but not OFF. In contrast, Ruitenberg et al. (19) investigating global proactive control (LWPCE) in a Stroop paradigm, found comparable conflict adaptation of movement speed in participants with PD both ON- and OFF-their dopamingeric medication.

Investigations regarding reactive control abilities in PD are sparse. To our knowledge, reactive control has only been assessed by Ruitenberg et al. (20) using the ISPCE. Controlling for S-R learning effects, Ruitenberg et al. (20) reported intact control adaptations in PD as compared to HC participants, independent of dopaminergic status.

Presently, the literature does not support a strong argument for the absence of local- and global proactive control in PD. The heterogeneous results, in conjunction with small sample sizes, of the reported results could suggest a potential reduction of local- and global proactive control in PD. Moreover, it has yet to be established that participants with PD are able to acquire the associated context-control rules independently of S-R learning in proactive control. Past studies did not distinguish between S-R associations effects and proactive control. While the latter has been done in one study on reactive control further corroborating evidence by replication is required.

### EEG Correlates of Adaptive Control

Electrophysiological recordings such as EEG have a high temporal resolution and are well suited to analyze cognitive and adaptive control predictions put forward by the DMC framework.

Electrophysiological correlates of cognitive- and adaptive control effects have often been investigated with event-related potentials (ERPs, for an overview see 18). Many of those ERPs have their spectral origin in the theta-band (4-8 Hz) (22). Not only the phase-locked part of theta oscillation (in other words the ERP), but to a larger degree also the non-phase locked part reflects conflict processing and is predictive of behavior (23). Theta modulation over midline-frontal electrodes are thought to communicate the need for control (22). Frontal theta has been shown to be reduced in people with PD in a number of processes/tasks, such as the startle-response (24) or interval timing (25). Moreover, with regard to conflict processing people with PD exhibit reduced response-related conflict activity at frontal electrodes relative to HC participants (26). Transient frontal theta activity has also been shown to increase in anticipation of a cognitive demanding task, indexing preparatory proactive control (27). Further, using a Simon task, Chinn et al. (28) found midline-frontal theta dynamics were modulated by proactive control adaptation (via LWPCE manipulation). In a temporal context where high conflict was expected (MI), less frontal theta was observed on conflict trials as compared to the low conflict temporal context (MC). Moreover, in local proactive control adaptation (as measured by the CSE) less frontal theta was observed when a conflict trial was preceded by a conflict trial (28,29). Pastötter et al. (29) traced the origin of the theta cluster of the CSE proactive control adaptation to the left cingulate gyrus and pre-supplementary motor area using multiple-source beamformer analysis. The opposite pattern can be expected for reactive control. Jiang et al. (30) in a Stroop-like task with an ISPCE manipulation, found activity in the theta band, in a posterior cluster, to be increased in incongruent items in high-conflict versus low-conflict conditions and congruent theta in congruent items to be increased in low-conflict conditions versus high conflict conditions. Thus, proactive control adaptation may be indexed by a reduction in control resources during conflict processing as indexed by a smaller midline-frontal conflict theta effect, whereas reactive control adaptation may be characterized by a larger conflict theta effect, possibly with a more posterior distribution.

### The Present Study

Given the heterogeneous results investigating adaptive control in PD in conjunction with methodological issues (such as sample size), the present study assesses if proactive and reactive control adaptation are affected in people with PD, using their usual medication regiment while controlling for S-R learning. We used a numerical Stroop paradigm (31), with only two response options and separate items to induce the manipulation (inducer items) and measure its effect (diagnostic items). Based on the available literature we expected reduced proactive control and intact reactive control in participants with PD in comparison to HC participants. Moreover, to distinguish reactive control effects from proactive control effects we further assessed midline-frontal theta activity. We hypothesized reduced midline-frontal theta band activity to be associated with successful adaptive control and impaired adaptive control to be indexed by a failure to adequately regulate midline-frontal conflict related theta activity. Moreover, we expect patterns in theta modulation to distinguish reactive from proactive control.

## Methods

### Ethics and Registration

The study was approved by the local medical ethics committee (2020-133) in accordance with the declaration of Helsinki (32). The study design was preregistered in the German clinical trial registry (DRKS00023020).

### Participants and General Procedure

We recruited 31 participants with PD and 34 healthy participants (HC) matched in age and gender. All PD patients fulfill the MDS-diagnostic criteria for PD (33). Four of the HC participants were excluded because they switched the response hand instead of using their right hand as instructed. One participant from the PD group had to be excluded as they were inattentive for large proportion of the task. The final data set included 30 participants (22 men and 8 women) in the PD group with a mean age of 64 years (SD = 9.6 years) and 30 participants (18 men and 12 women) in the control group with a mean age of 59.4 years (SD = 6.8 years).

We only included participants who fulfilled the following criteria: a) no co-morbid neurological or psychiatric problems, b) right-handedness, c) normal or corrected to normal vision, d) MMSE <25 (mean_PD_ = 29.2, SD_PD_ = 1.1; mean_HC_ = 29.1, SD_HC_ = 1.0). UPDRS-III ratings were recorded for all participants with PD (mean = 12.0, SD = 7.0). Moreover, we recorded the formal education in years of our participants (mean_PD_ = 16.2 y, SD_PD_ = 3.0 y; mean_HC_ = 16.0 y, SD_HC_ = 3.2 y).

The experiment took part at the out-patient clinic of the Evangelisches Krankenhaus Oldenburg. Since the recordings took part around the beginning of the Covid-19 pandemic, participants and experimenters remained masked for the duration of the experiment. After being informed about the purpose of the experiment, participants gave their written informed consent and were screened with the MMSE and reported their education level in years. MDS-UPDRS-III ratings were recorded for all participants with PD. Subsequently, participants performed the experiment and EEG was recorded. In total, task and preparation lasted for about 2.5 hours.

### Experimental Paradigm

Participants performed a numerical Stroop task with both LWPCE and ISPCE manipulations, presented in OpenSesame (34). The numerical Stroop task requires participants to select the numerically larger or smaller number between two numbers displayed on screen. Stroop-like congruency-conflict effects are introduced by manipulating the physical size of the displayed number pairs. One number was always numerically larger to the other and one number was displayed physically larger than the other.

For example, in an incongruent comparison the numerically smaller number is displayed physically larger and the numerically larger number is displayed physically smaller. Our rationale for selecting the numerical Stroop task was twofold: 1) to have sufficiently many items available to have inducer and diagnostic item sets, and 2) reduce task demands and avoid for our participants having to learn multiple response options (e.g. color associated keys in the Stroop task). During each trial participants saw a fixation cross for 300-600ms (uniformly varied), followed by the two numbers (for a list of items see Table 1) presented on screen until participants indicated a response. Participants had up to 2000ms to make aresponse. Afterwards, a blank screen was displayed for 800ms before the next trial started.

**Table 1.** List of items used in the numerical Stroop task. We used items with a numerical distance 1 and 2 also previously used by Dadon and Henik (35). Items were balanced in terms of numerical presentation and overall presentation. For ISPCE manipulation either small or large number pairs were manipulated respectively.

| Items |  |  |
| --- | --- | --- |
| Numerical Distance | Small Number Pairs | Large Number Pairs |
| 1 | 1-2, 3-4 | 6-7, 8-9 |
| 2 | 1-3, 2-4 | 6-8, 7-9 |

Participants completed a total of 8 blocks with 134 trials each. Four blocks contained the LWPCE items and the other four blocks contained the ISPC items. Per Block ~70% of the items presented were inducer items (N = 94) and ~30% (N = 40) were diagnostic items. Diagnostic items contained equal proportions of congruent and incongruent items. Inducer items contained ~80% incongruent items in the MI condition and 80% congruent items in the MC condition. In the LWPCE manipulation, participants were presented with blocks containing more- (MI - 70% incongruent- 30% congruent trials) and two blocks containing fewer (MC - 30% incongruent- 70% congruent trials) incongruent items. In each of the four ISPCE blocks equal amounts of congruent and incongruent items (see Figure 1) were presented. Only the proportions of half of the items were biased. We chose to manipulate the proportion of congruency by the numerical size of the number pairs - larger number pairs (numbers > 5) versus smaller number pairs (numbers < 5; see Table 1). For example, two large number pairs were randomly selected as inducer items, and the remaining two large number items were subsequently used as diagnostic items. For half of the participants (per experimental group) the large number items contained MC comparisons and the small items MI comparisons and vice versa for the other half of the participants. Before performing the task, participants completed 32 trials with feedback, without the LWPCE or IWPCE manipulations, to familiarize themselves with the task.

**Figure 1.**
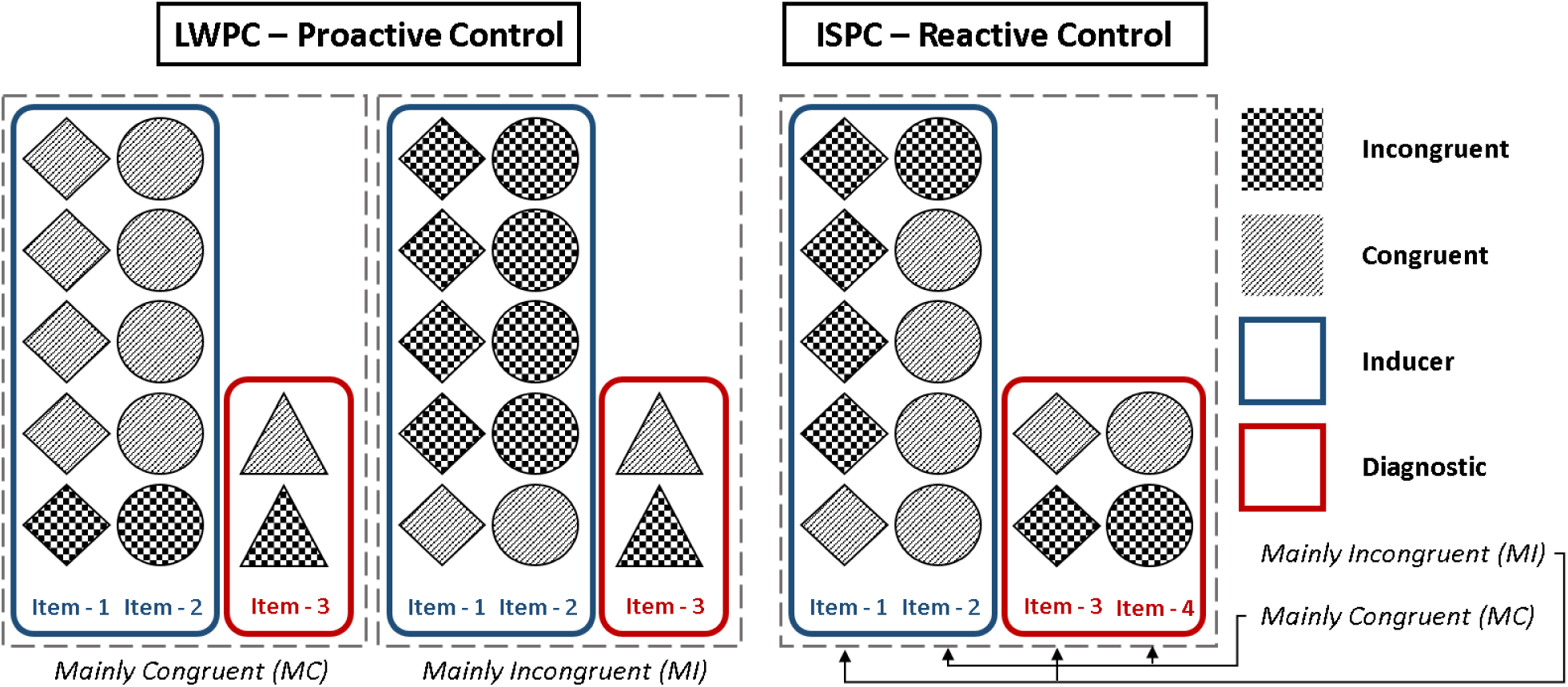
Schematic presentation of the task design. Proactive control was manipulated via the LWPCE with blocks consisting of either mainly congruent (MC) and mainly incongruent (MI) inducer items (blue). Reactive control vice versa with mainly congruent and incongruent inducer items, the manipulation was induced per item (large versus small number pairs). Effects were measured in unbiased diagnostic items (red).

Block presentation order was randomized and participants would either start with the ISPCE blocks or the LWPCE blocks. Within the LWPCE blocks we also randomized whether the MI or MC block was presented first. Moreover, items were presented to the participants in pseudo-randomized order. Pseudo-randomization was performed with custom-written Python scripts, implementing the algorithm by van Casteren and Davis (36), using a mix of random shuffling and backtracking to achieve a random presentation of the criteria we defined. Criteria were (1) diagnostic items should not be presented one after each other (in order to have a balanced presentation of diagnostics throughout the block) (2) the same item should not be presented more than twice in a row, (3) inducer items should not be presented more than four times in a row, (4) the same correct response side (left or right) should not be presented on more than three consecutive trials and (5) the same congruence should not follow be presented more than four times in a row. Item presentation order were randomized for each participant separately according to these rules.

### Behavioral Data Analysis

Analysis of the behavioral data was performed in R (R-4.1.3) (37). Data visualizations were created using the ggplot2 package (38) and ggdist (39).

Statistical analyses of the behavioral data were performed using Bayesian mixed effect models using the brms package (40). For the RT analysis (performed on correctly answered trials) due to right-skewed distribution of RT data we used a shifted log-normal likelihood function. Trials shorter than 200ms were not included in the analysis. In total 20 trials or 0.03% of the data were excluded. The ISPCE and LWPCE were analyzed in separate models. We used contrast coded dummy variables to calculate the main effects for Congruency (congruent - incongruent), Block/Item Type (MC - MI) and their interaction for each group (PD - HC) separately (see supplement – section 1). Moreover, we included random intercepts for the participants and items. Further, we included separate intercepts per group for the shift parameter of the model.

In order to maximize the utility of the data we collected, we used information from the inducer items to inform the prior parameter space of the diagnostic models. We fitted separate inducer models for the LWPCE- and ISPCE data. We used weakly informative priors derived at by prior predictive simulations yielding plausible RT distributions of the data (see supplement – section 2). For the diagnostic model we used the posterior distribution of the inducer model to construct informed priors for the effects not expected to differ between conditions (shift parameter, sigma, Congruency, Block/Item, and the random intercepts). We used a Gaussian distribution, with mu equal to the mean of the inducer posterior and sigma defined as the larger absolute difference, between the posterior mean and the two 95% credible interval borders of the inducer posterior. For the interaction effects we used regularized priors with a Gaussian distribution centered around zero with sigma defined as the larger absolute difference, between zero and the two 95% credible interval borders of the posterior distribution of the inducer model.

To evaluate the interaction effect between Congruency and Block/Item (depending on ISPCE or LWPCE analysis) in milliseconds, we used the posterior distribution to calculate the estimated marginal means for each effect (MC congruent, MC incongruent, MI congruent, MI incongruent) and group. Next, we calculated the “Conflict effect” (congruent - incongruent) for each of the two conflict conditions (MC and MI). Adaptive control was operationalized as the difference between the two conflict effects. Moreover, we compared how RT varied in congruent and incongruent trials between the two conflict conditions (i.e. MI congruent - MC congruent and MI incongruent - MC incongruent).

For the analysis of the error data we performed a logistic regression (Bernoulli-distribution with logit-link function) and used the same approach to fit inducer and diagnostic models as outlined for the shifted-log normal model. Priors were weakly informed so that lower error rates had a higher probability, as typically participants never make more than 10% errors during these kinds of tasks data (see supplement – section 3). Log-odds of the estimated marginal mean effects were transformed into probabilities for a more meaningful interpretation.

To evaluate how well a particular model factor predicted the data we calculated Bayes inclusion factors (BIF) across matched models using bridge sampling (41). We calculated the likelihood for the factors of each experimental group separately. For example, to evaluate the contribution of Congruency in group HC, we did not consider the contribution of any of the factors in the PD group. The BIF of Congruency reflects the likelihood of factor Congruency (*H_A_*) over the averaged likelihood of the null model and the model containing the factor Block (*H*_0_). BIFs and BFs smaller than 1 suggest evidence in favor of the null model (no contribution of the factor/interaction in question), whereas BIFs and BFs larger than 1 would signify evidence in favor of the alternative. We qualify evidential strength provided by the BIF by using criteria suggested by Jeffreys (42). BIFs smaller than 3 and larger than 1/3 provide insufficient evidence for either hypothesis. BIFs between 3 and 10 (or 1/3 and 1/10) provide anecdotal evidence, between 10 and 30 (1/10 and 1/30) strong evidence and anything larger than 30 (or < 1/30) is classified as substantial evidence in support of the hypothesis in question.

For the Markov chain Monte Carlo (MCMC) sampling we ran four chains with 2000 warm-up iterations and 10000 iterations for each chain to sample from the posterior distribution. We assessed model convergence by confirming that the potential scale reduction factor *R̀* for all parameter were near 1 and less than 1.1 and by visual inspection of the chain trace plots. Model fit was assessed comparing the actual data with simulated data from the model’s posterior predictive distribution (see supplement section 5).

#### Exploratory Analysis

After identifying a deficit for the LWPCE manipulation in the PD group, we were interested in the association between proactive control and motor status of the participants. We fitted the inducer and diagnostic model (with the previously informed priors) to the data of the participants with PD alone. Again, we assessed effects of Block, Congruency and the interaction. In contrast to the previous models we included random slopes for the main and interaction effects. The resulting model gave us participant specific posterior distributions. We calculated proactive control as the difference in Stroop effects between the two conflict conditions (MC and MI), with a posterior distribution of the differences for each participant. We summarized proactive control as the mean of the resulting posterior distribution and calculated the Pearson correlation with the participant associated MDS-UPDRS motor scores. To evaluate statistical significance in this analysis we calculated p-values with a significance level alpha of 0.05.

### EEG Recording and Preprocessing

EEG was recorded continuously (actiChamp plus by Brain Vision 43) with a sampling rate of 1000 Hz, from 128 active Ag/AgCl electrodes. Impedances were kept below 10kΩ. Preprocessing was performed in Matlab using the EEGLAB toolbox (44). Data were first downsampled to 250 Hz, 0.1 Hz high-pass filtered and detrended over the whole recording period for each channel separately. Noisy channels were excluded via the pop_clean_rawdata() function with the following criteria: FlatlineCriterion = 5, ChannelCriterion = 0.8, and LineNoiseCriterion = 5. On average we identified and removed 4.9 (SD = 4.1) bad channels per participant. Afterwards, data were re-referenced to an average reference. Line-noise was removed using EEGLABs pop_cleanline() function. Noisy data segments were excluded in a automatized manner with artifact subspace reconstruction (45) using the pop_clean_rawdata() function with the “Burstcriterion” parameter set to 80. We selected a relatively high threshold here in order to remove only excessively noisy data segments (e.g. movement occurred during the segment in question) and not eye blinks. Subsequently we used independent component analysis (Infomax algorithm) to identify and remove components reflecting eye blinks and larger muscle artifacts. Data were then epoched stimulus-locked [−1.2s, 2s] and response locked [−2s, 1.2s]. In order to decrease the effects of volume conduction, we performed a surface-Laplacian transformation using the CSD toolbox (46).

After preprocessing in the group with PD, we had to exclude one participant due to insufficient data quality after preprocessing (due to excessive movement during recording), and for two participants after preprocessing there were too few trials in the ISPCE task in some conditions (<40) to perform the planned regression analysis. The same was true for two participants in the LWPCE condition. Thus, the final data set in the LWPCE and ISPCE analyses of the PD group (including the participant excluded due to poor behavioral performance) consisted of 26 participants. In the HC group we could not use the data of one participant due to a technical problem during recording and had too few trials (<40 per condition) for the regression analysis in one participant in the ISPCE analysis. The final data set of the HC group consisted of 29 participants for LWPCE analysis and 28 participants in the ISPCE analysis.

### EEG Data Analysis

Response-locked and stimulus-locked data sets were frequency transformed via fast Fourier transform (FFT) and convolved with the FFTs of a series of Morlet wavelets. We used 20 wavelets logarithmically spaced from 2 Hz to 30 Hz. Wavelet cycles were logarithmically spaced from three to ten, increasing by frequency. This allowed better band specific resolution for the higher frequencies and better time resolution for lower frequencies. After wavelet convolution data was down-sampled to 125Hz for further analysis. Due to a short fixation window (600-300 ms) baseline correction was performed over the whole trial period (47). A baseline period covering 0 ms to 1000 ms (or −1000 ms to 0 ms for the response locked data) was used for decibel conversion. This approach allowed us to identify transient changes in oscillatory activity.

After decibel conversion, we performed a GLM analysis using the ordinary least-squared solution for each participant, electrode, frequency and time point. To calculate the regression coefficients, we used functions provided by the LIMO toolbox (48). For the GLM of the EEG data, we used similar effect coded contrasts as in the behavioral analysis, with separate models for the LWPCE and ISPCE data. For each participant we had effect coded contrast for Congruency, Block/Item and their interaction. We did not distinguish between inducer and diagnostic items, as the number of trials in the diagnostic items alone, after preprocessing would have been too low. As with the behavioral data, to investigate the interaction effect between Block/Item and Congruency, marginal mean effects were calculated for each participant separately. With the marginal mean effects, we calculated the difference in Congruency (Congruent - Incongruent) for the conflict effect in each block/item (MC and MI). Adaptive control (for the ISPCE and LWPCE separately) was then analyzed by comparing the difference in conflict effects in blocks or items when proportionally more conflict is expected versus when proportionally little conflict is expected (MI - MC). To investigate the contribution of conflict-related theta activity, we then averaged the data over the theta-band (4-8 Hz). In order to correct for multiple comparisons, we used non-parametric cluster-based permutation test statistics (49) in FieldTrip (50) using dependent t-tests, a cluster-alpha of 0.05 and an alpha of 0.05, with 10000 permutations.

## Results

### Behavioral Results

Overall, we found insufficient evidence that participants with PD were slower (747 ms) compared to the HC participants (673 ms) in overall RT performance, *t*(58) = 2.11, *p* = 0.039, *BF_10_* = 1.66 (see Figure 3). We found strong evidence that participants with PD performed on average more errors (3.1%) than the HC participants (1.5%), *t*(58) = −3.1, *p* = 0.004, *BF_10_* = 11.53. Nonetheless, overall accuracy in the task was high across groups and we found only very limited evidence for the use adaptive control in the analysis of the error data (the interested reader is referred to section 4 in the supplement, for a summary of the results).

The results of the shifted-log normal analysis are depicted in Table 2 and Figure 2. With regard to adaptive control manipulations, the RT analysis of the inducer items of the LWPCE manipulation showed an interaction effect, with reduced conflict RT in the MI versus MC condition, for both the HC participants (*m* = −47.8 ms, *CI* = (−62.1 ms| −34.3 ms), *BIF* > 1000) and the participants with PD (*m* = −35.9 ms, *CI* = (−49.6 | −22.6 ms), *BIF* > 1000). The effect in the HC group was explained to a large degree by a reduction in RT to incongruent items in the MI condition (*m* = −29.9 ms, *CI* = (−40.5 ms | −19.8 ms)) and to a lesser degree by a reduction of RT to congruent items in the MC condition (*m* = −17.9 ms, *CI* = (−26.6 ms | −9.6 ms)). The PD group showed a similar pattern with a strong reduction in RT to incongruent items in the MI condition (*m* = −26.2 ms, *CI* = (−36.8 ms | −16.2 ms)) and slightly less reduction of RT in congruent items in the MC condition (*m* = −9.6 ms, *CI* = (−18.3 ms | −1.3 ms)). In the analysis of the diagnostic items, we also found decisive evidence for the presence of an interaction effect in the HC group, (*m* = −34.5 ms, *CI* = (−51.5 ms| −18.2 ms), *BIF* = 518.32), with the effect being driven by a reduction in RT to incongruent items in the MI condition (*m* = −27.8 ms, *CI* = (−39.9 ms | −16.3 ms)) but possibly no reduction of RT to congruent items in the MC condition (*m* = −6.7 ms, *CI* = (−16.5 ms | 2.8 ms)), as the credible interval was compatible with null. The interaction effect in the diagnostic items was smaller in the PD sample and provided insufficient evidence (*m* = −17.2 ms, *CI* = (−33.7 ms| −1.1 ms, *BIF* = 1.74)), for the presence of interaction effect. Contrasts revealed a relative reduction in RT congruent items in the MC condition (*m* = −10.1 ms, *CI* = (−19.9 ms| −0.3 ms)) but possibly no reduction in RT to incongruent items in the MI condition (*m* = −7 ms, *CI* = (−18.5 ms| 4.3 ms)) with the credible intervall being compatible with null. Moreover, the difference between groups in posterior probability in RT reduction to incongruent items in the MI condition indicates that HC participants had a larger reduction with a mean of 20.8 ms and 95% of the posterior probability between 37.1 ms and 4.8 ms. Thus, it appears that relative to HC participants, participants with PD were impaired in their ability extend proactive control to diagnostic items. Figure 3B also highlights this qualitative difference between HC and participants with PD. The RT of the diagnostics items tracks the RT of the inducer items closely for the HC participants over time, but not the participants with PD.

**Figure 2.**
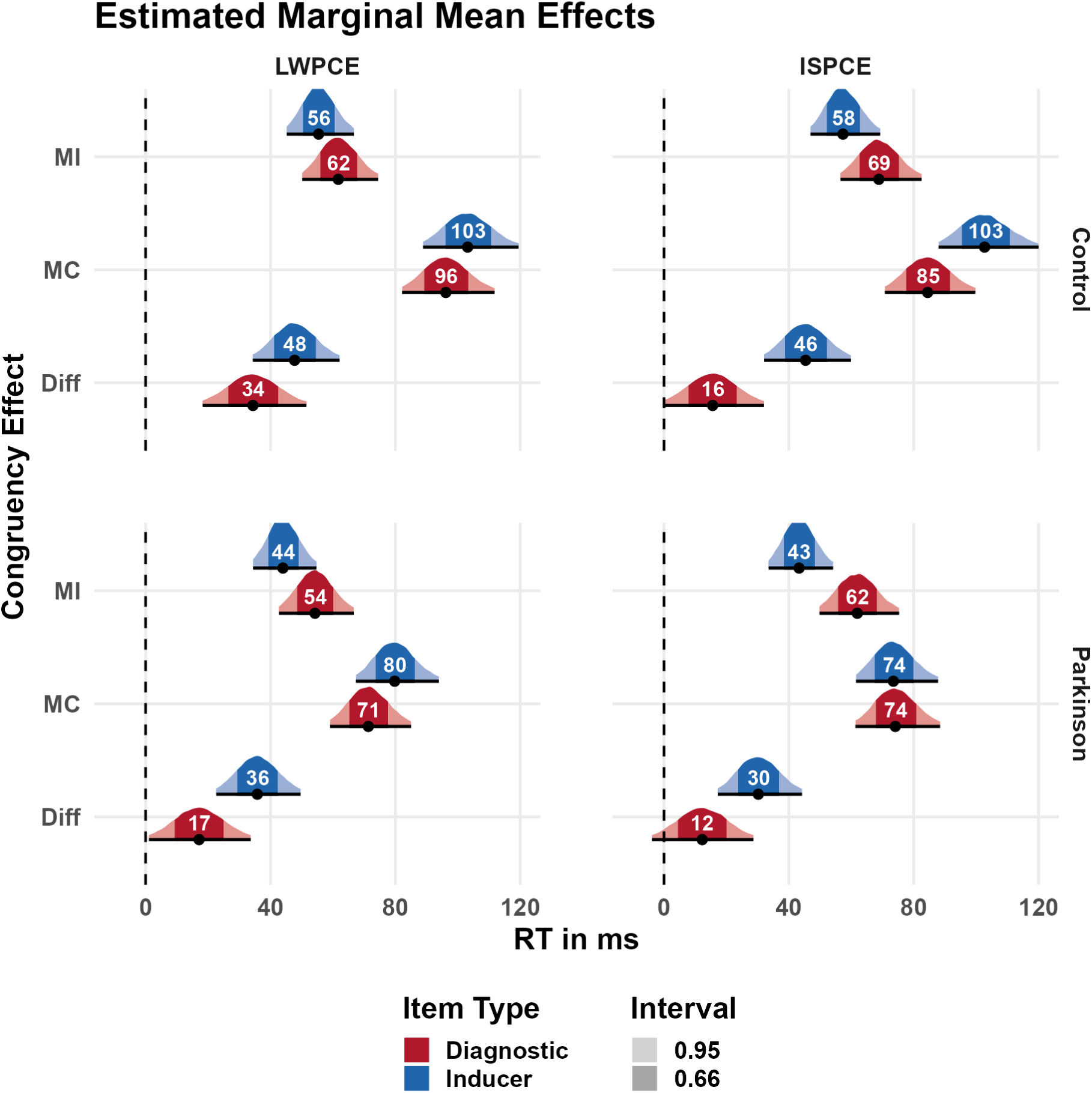
Summary of the shifted log-normal regression analysis results. Displayed are the estimated marginal mean posterior distributions for the conflict effects (incongruent - congruent) in the high conflict condition (MI), low conflict condition (MC) and their difference. Results are organized row-wise by group (Control and Parkinson) and column-wise by effect (LWPCE and ISPCE). Inducer model estimates are displayed in blue and diagnostic model estimates in red. The shaded areas of the posterior distribution correspond to the 66% and 95% credible intervals.

**Figure 3.**
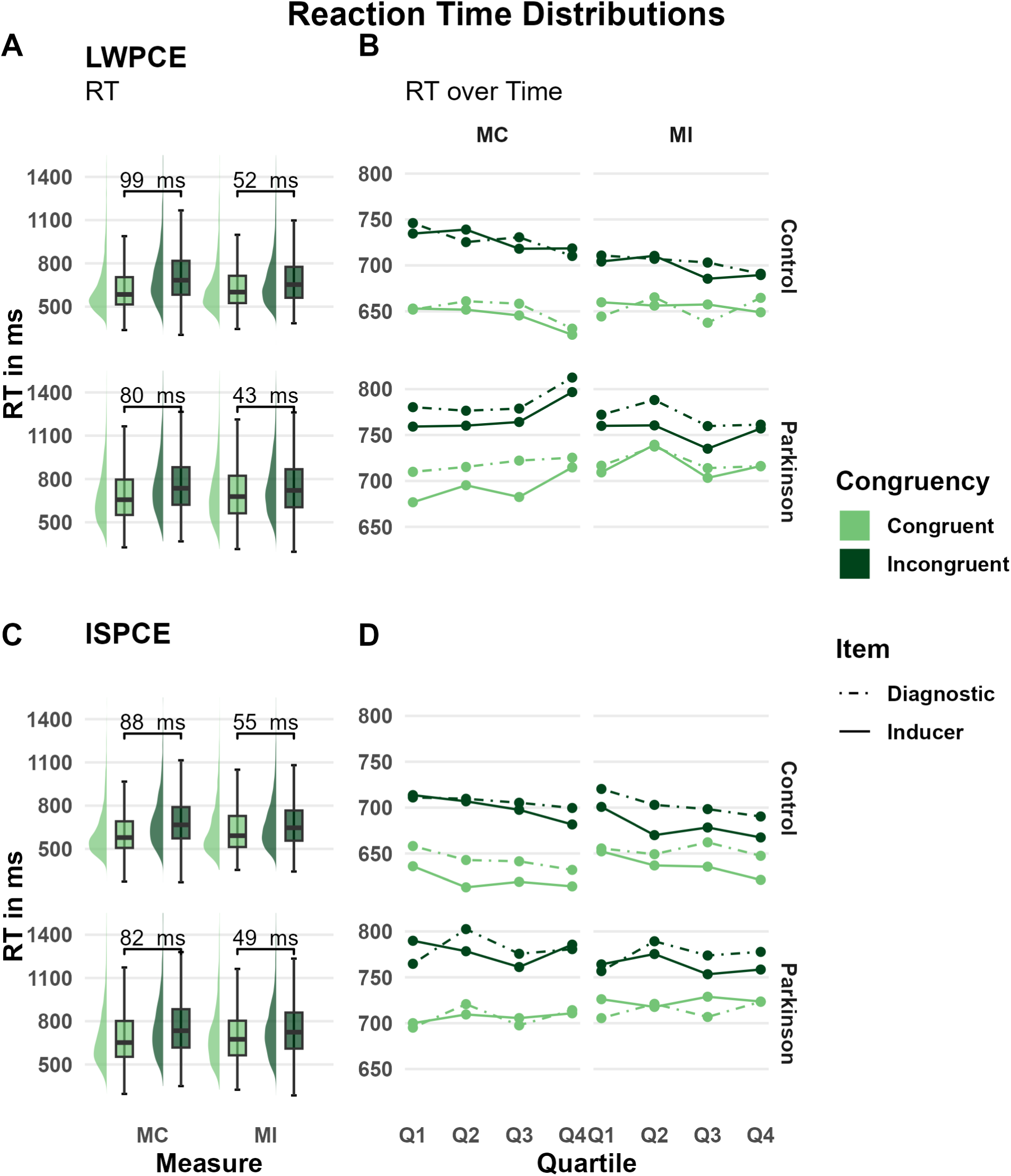
Reaction time distributions and consistency of diagnostic and inducer item reaction times over time of the LWPCE- (A, B) and ISPCE data (C, D). The top row contains data for the HC participants and bottom row of the participants with PD. Panel A) and C), depict the RT distribution of all items (diagnostic and inducer) by congruence for the high conflict (MI) and low conflict (MC) proportion manipulation. Nodes at the bottom show the average difference in RT between congruent and incongruent item by proportion manipulation. Panel B) and D) display the time course of RT averaged over quartiles by congruency (congruent and incongruent) and item type (inducer and diagnostic items). The two columns distinguish the high- (MI) and low (MC) conflict proportion manipulation.

**Table 2.** Results of the shifted log-normal regression analysis. Mean estimates and 95 percent credible intervals are provided in milliseconds. The factor congruency reflects the difference between incongruent and congruent items, the factor Block/Item PC the relative difference between MC and MI blocks/items and the interaction reflects the difference in conflict effects (incongruent - congruent) in the MI blocks/items relative to the MC blocks/items. BIF exceeding 1000 or smaller 0.001 are abbreviated for the purpose of making the table legible.

| Parameter | PD |  | HC |  |
| --- | --- | --- | --- | --- |
| | $M(CI)$ in ms | $BIF_{10}$ | $M(CI)$ in ms | $BIF_{10}$ |
| **LWPCE – Inducer Items** |  |  |  |  |
| Congruency | 62.1(52.9/72.3) | >1000 | 79.5(68.7/91.4) | >1000 |
| Block PC | 8.3(1.8/14.9) | 0.19 | 6.0(-0.5/12.6) | 0.04 |
| Interaction | -35.9(-49.6 / -22.6) | >1000 | -47.8(-62.1/-34.3) | >1000 |
| **LWPCE – Diagnostic Items** |  |  |  |  |
| Congruency | 62.9(53.9/72.7) | >1000 | 79.1(68.9/90.4) | >1000 |
| Block PC | -1.5(-8.4/5.3) | 0.35 | 10.6(3.7/17.7) | 14.80 |
| Interaction | -17.2(-33.4/-1.1) | 1.74 | -34.5(-51.5/-18.2) | 518.32 |
| **ISPCE – Inducer Items** |  |  |  |  |
| Congruency | 58.6(49.5/69.0) | >1000 | 80.3(69.2/93.0) | >1000 |
| Item PC | -1.1(-7.4/5.5) | 0.03 | 20.9(14.1/28.1) | >1000 |
| Interaction | -30.4(-44.2/-17.3) | >1000 | -45.5(-59.9/-32.1) | >1000 |
| **ISPCE – Diagnostic Items** |  |  |  |  |
| Congruency | 68.2(58.3/79.1) | >1000 | 76.9(66.2/88.5) | >1000 |
| Item PC | -6.2(-13.3/0.9) | 3.00 | 17.0(9.8/24.4) | 235.78 |
| Interaction | -12.3(-28.7/3.9) | 0.68 | -15.6(-32.0/0.2) | 0.53 |

The analysis of the inducer items of the ISPCE analysis revealed again decisive evidence for the presence of an interaction effect in both the HC- (*m* = −45.5 ms, *CI* = (−59.9 ms | −32.1 ms), *BIF* > 1000) and PD participant (*m* = −30.4 ms, *CI* = (−44.2 ms | −17.3 ms), *BIF* > 1000) groups. The effect in the HC group was determined by a RT reduction to the incongruent items in the MI condition (*m* = −43.6 ms, *CI* = (−55.3 ms | −32.8 ms)) but possibly no reduction in RT to congruent item in the MC condition (*m* = −1.9 ms, *CI* = (−10.1 ms | 6.2 ms)) with null being included in the 95% credible interval. The interaction effect in the group with PD, was explained by a relative decrease in RTs of congruent items in MC condition (*m* = −16.2 ms, *CI* = (−25.1 ms | −7.6 ms)) and a reduction in RTs of incongruent trials in the MI condition (*m* = −14.2 ms, *CI* = (−24.3 ms | −4.6 ms)). Comparing the difference between groups in posterior probability in RT reduction to incongruent items in the MI condition showed that HC participants had a larger reduction with a mean of 24.8 ms and 95% of the posterior probability between 41.6 ms and 8.6 ms. Thus, while both groups show a sizable ISPCE effect on RTs in the inducer items, these appear to have different origins. The HC participants appeared to be more effective in regulating responses to conflict relative to the participants with PD. Further analysis of the diagnostic items provided no evidence for an ISPCE in either group and instead anecdotal evidence favoring the null hypothesis in the HC- (*m* = −15.6 ms, *CI* = (−32.0 ms | 0.2 ms), *BIF* = 0.53) and group with PD (*m* = 12.3 ms, *CI* = (−28.7 ms | 3.9 ms), *BIF* = 0.68). Hence, we found no evidence that either group could extend reactive control beyond the items used to induce the ISPCE manipulation. The mismatch between averaged diagnostic- and inducer RT in both groups is highlighted in **Error! Reference source not found.**B.

#### Exploratory Analysis - Symptoms

There was no statistically significant correlation between MDS UPDRS motor scores and LWPCE in the inducer items (*r* = 0.35, *p* = 0.067), and the diagnostic items (*r* = 0.28, *p* = 0.14).

#### EEG Results

We assessed whether conflict related power in the theta band was reduced in context when high conflict is expected (MI) versus contexts when little conflict is expected (MC). Figure 4A and Figure 4B plot the conflict effect (incongruent - congruent) at the FCz channel for LWPCE. As expected, power appears to be larger, at frequencies between 4-8 Hz, on incongruent trials than congruent trials (condition specific plots of the items can be found in section 6 of the supplement). Moreover, it appears that the conflict effect is larger in the MC condition, than the MI condition particularly in the HC group. To evaluate the difference in conflict effects between conflict context manipulations further we used non-parametric, cluster-based permutation testing. The HC participants showed a significant negative cluster after stimulus presentation (0.35 s to 0.77 s; *p* = 0.0004), largely over midline-frontal to frontal-right channels. A negative cluster could also be observed prior to action onset (−0.6 s to −0.05 s; *p* = 0.002), with a more midline-frontal distribution. Thus, both the response-locked (RL) and stimulus-locked (SL) data indicate reduced conflict midline-frontal theta when more conflict is expected. Looking at the same analysis in the PD group we also find significant negative clusters after stimulus presentation (0.33 s to 0.71 s; *p* = 0.0088) and prior to response onset (−0.6 s to −0.14 s; *p* = 0.0012). Both clusters show a frontal left distribution (see figure Figure 4). Both groups show a reduction in midline-frontal conflict related theta activity when more conflict is expected. Figure 4E and Figure 4F further show the modulation by estimated marginal mean condition averaged over the electrodes identified in the significant cluster. For the HC participants it appears that conflict expectancy modulates both theta levels of congruent and incongruent trials, whereas in the PD group only incongruent trials appear to be reduced.

**Figure 4.**
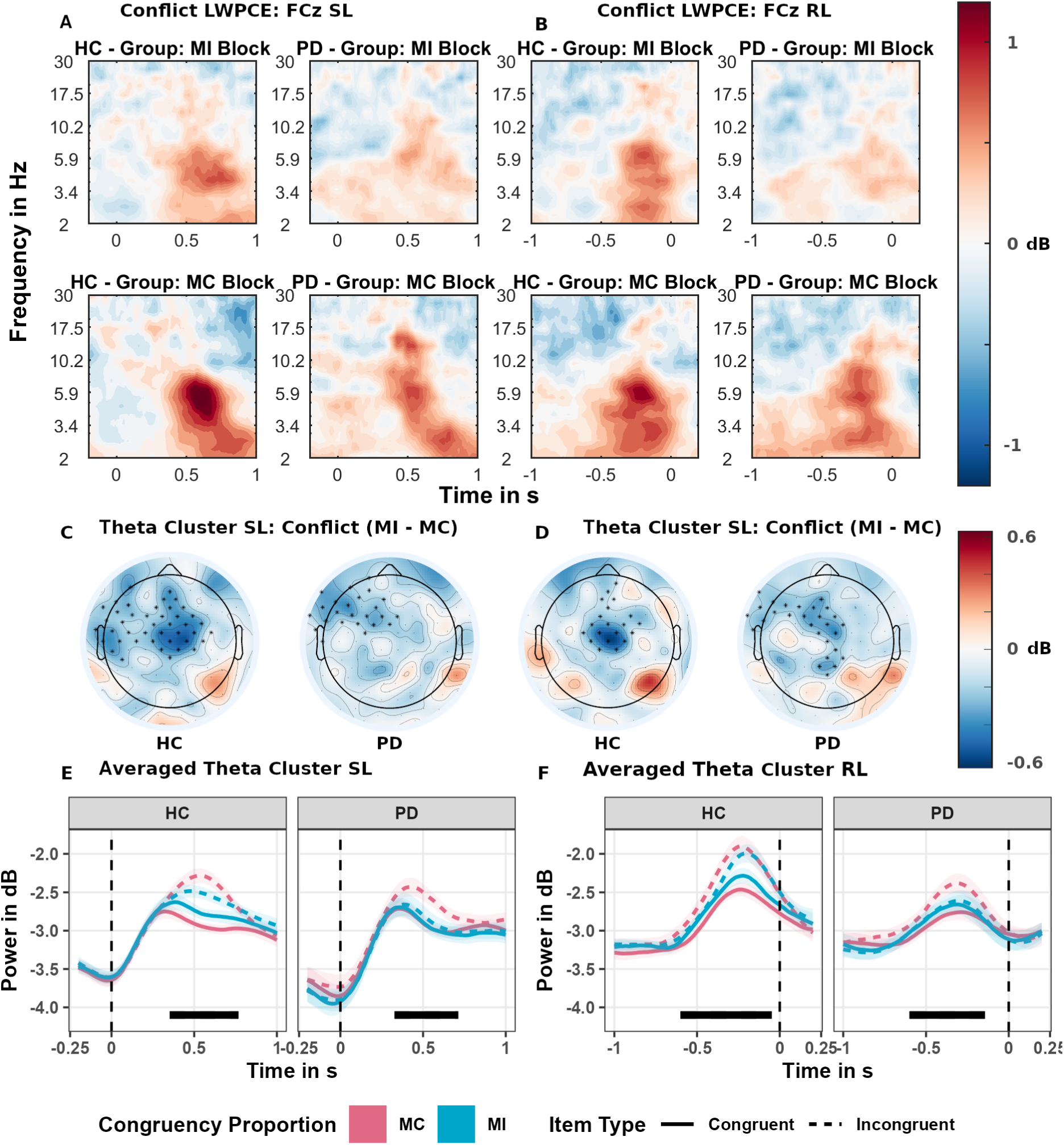
Estimated marginal mean effects of the time-frequency regression analysis for the LWPCE manipulation. Data on the left (A, C, E) are plotted in reference to stimulus onset (SL) and data on the right (B, D, F) are referenced to the response (RL). Panels A and B show the time-frequency results at the FCz electrode (A, B) for the conflict effect (incongruent - congruent) by conflict proportion manipulation (MI and MC) and group (HC and PD). Panels C and D depict the results of the theta band (4-8 Hz) cluster-permutation analysis on the difference in conflict effects between conflict proportion manipulation (MI - MC). Significant clusters are marked with an asterisk symbol. The underlying color gradient depicts the averaged difference in conflict theta activity over the period when the significant cluster was detected. Panels E and F depict the estimated marginal mean effects in theta activity, averaged over electrodes of the significant cluster. Shaded areas depict the standard errors (SE) at each sample. Marked in black is the period when the significant cluster was detected.

For the ISPCE Figure 5A and Figure 5B similarly as for the LWPCE it appears that the conflict effect at the FCz electrode is larger, at frequencies between 4-8 Hz, in the MC condition, than the MI condition in the HC group. Further, cluster-based permutation tests only the HC participants showed significant differences in conflict related theta. A negative cluster was observed over midline-frontal electrodes (see Figure 5C and Figure 5D) both post stimulus onset (0.39s to 0.75s; *p* = 0.0017) and prior to response (−0.44 s to −0.1s; *p* = 0.0039). Thus, only the HC participant showed reduced conflict under conditions where high conflict was expected with certain items, as compared to when low conflict was associated with certain items. Estimated marginal means averaged over the significant cluster in Figure 5E and Figure 5F it appears that in the HC participants conflict expectancy modulates theta levels incongruent trials but not congruent trials.

**Figure 5.**
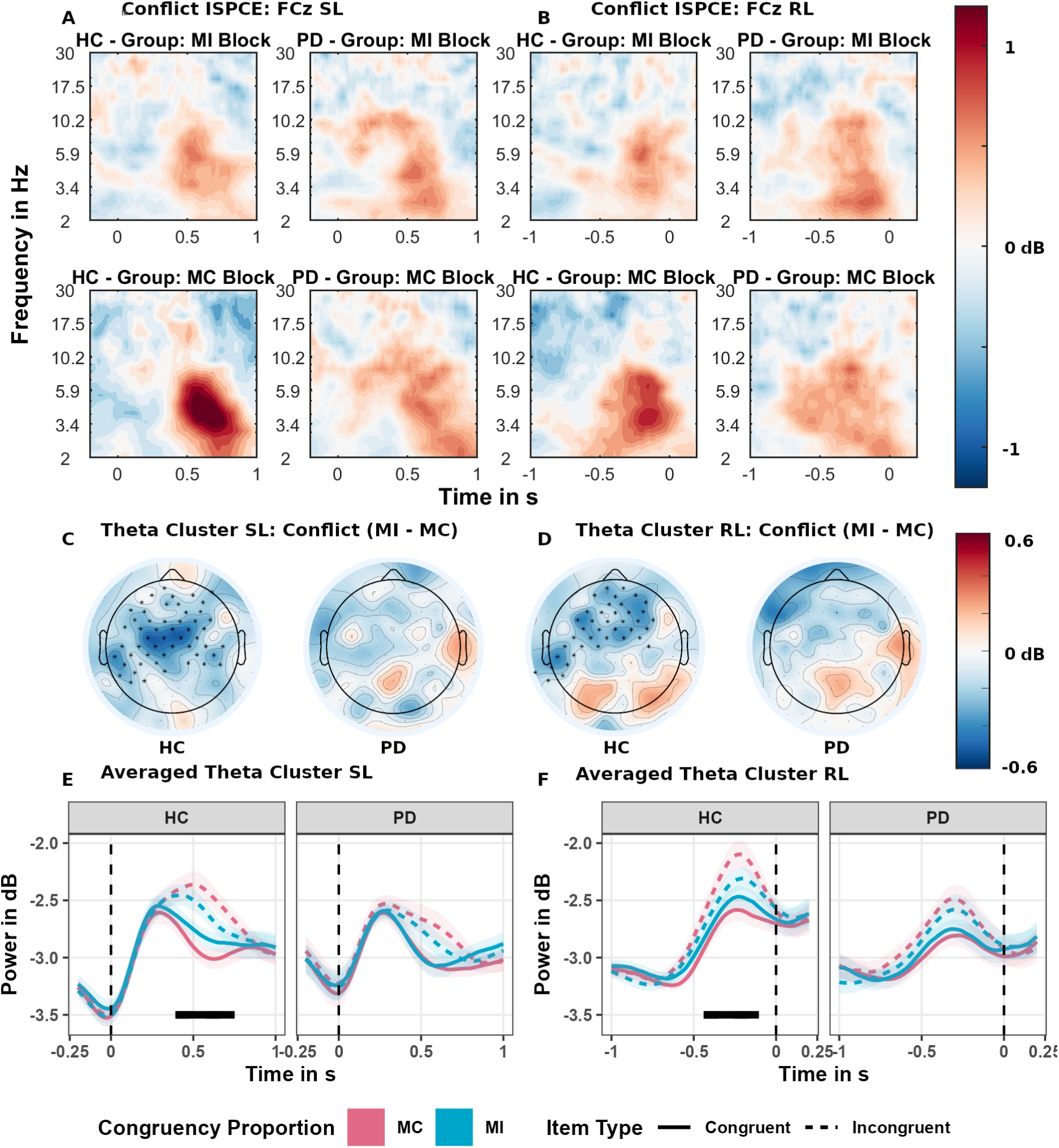
Estimated marginal mean effects of the time-frequency regression analysis for the ISPCE manipulation. Data on the left (A, C, E) are plotted in reference to stimulus onset (SL) and data on the right (B, D, F) are referenced to the response (RL). Panels A and B show the time-frequency results at channel FCz (A, B) for the conflict effect (incongruent - congruent) by conflict proportion manipulation (MI and MC) and group (HC and PD). Panels C and D depict the results of the theta band (4-8 Hz) cluster-permutation analysis on the difference in conflict effects between conflict proportion manipulation (MI - MC). Significant clusters are marked with an asterisk symbol. The underlying color gradient depicts the averaged difference in conflict theta activity over the period when the significant cluster was detected. In the PD panels where no significant difference was detected, we display the activity between 0.3s and 0.7s and −0.6s and −0.2s for comparison. Panels E and F depict the estimated marginal mean effects in theta activity, averaged over electrodes of the significant cluster in the HC group. Shaded areas depict the standard errors (SE) at each sample. Marked in black is the period when the significant cluster was detected.

## Discussion

In the present study we sought to investigate whether adaptive control is impaired in medicated participants with PD. To this end, we created a task to investigate both proactive control- and reactive control adaptation and compared performance in participants with PD to HC participants. To avoid confounding S-R learning we distinguished between manipulation inducing item and unbiased diagnostic items. Our results show an impairment in the acquisition of general context-control associations for proactive control adaptation in PD. Further, our results also suggest a partial impairment in reactive control, as participants with PD showed evidence of reactive conflict adaptation, but in contrast to HC participants, PD individuals were unable to improve performance on conflict trials.

### Interpretation Adaptive Control in PD

We observed a specific impairment to form general temporal context-control associations by participants with PD in the task requiring proactive control. The initial analysis of the inducer RT data showed strong evidence for the ability of both PD and HC participant to use proactive control. Both groups displayed reduced RTs on incongruent trials, in high conflict temporal context (MI), compared to low-conflict temporal context (MI). However, in the assessment of the unbiased diagnostic items we observed strong evidence that only the HC-, but not the participants with PD, were able to regulate proactive control. This qualitative difference between both groups in the diagnostic data demonstrates that participants with PD are impaired in the ability to learn context-control associations necessary for proactive control.

Our findings on the inducer items are in line with Ruitenberg et al. (19) who observed significant conflict adaptation effects in participants with PD both ON- and OFF-their dopaminergic medication performing a Stroop task. Our diagnostic manipulation extends results by Ruitenberg et al. (19) showing that this adaptation effect does not transfer to general context dependent adaptation, but is restricted to the items which induce it. However, our results are in contrast to Bonnin et al. (6) reporting evidence for proactive, global control adaptations in healthy control participants, but not participants with PD (DOPA-ON). The insignificant interaction effect in participants with PD was interpreted as evidence for the null hypothesis of impaired conflict modulation. However, Bonnin et al. (6) never directly compared proactive control in participants with PD to HC participants. Similar to Bonnin et al. (6) our post-hoc analysis did not show an association with UPDRS scores. Thus, the impaired ability to extend control to a broader context, may not be associated with disease progression and could present a general deficit, or one that is present early on.

Behavioral results in conjunction with the absence of electrophysiological evidence suggest that participants with PD may also show impairments in reactive control adaptation. Both participants with PD, and HC participants displayed strong evidence for an interaction effect, indicative of reactive control in the analysis of the inducer items. However, inspection of the congruent and incongruent trials by conflict context revealed that only HC participants were effectively able to reduce RT conflict cost on incongruent trials in items associated with high conflict. The interaction effect in participants with PD on the other hand was largely explained by facilitation effects on congruent items in low conflict context items. The subsequent analysis of the diagnostic items showed no evidence for reactive control adaptations in both groups. This could be due to a failure of our item manipulation which will be discussed subsequently. However, since in the group with PD a reduction in conflict cost in the high conflict items was absent in the inducer items, a carry-over effect to the diagnostic items is unlikely. This could indicate that participants with PD are able to shift attention toward specific features to a degree but may still be unable to exert reactive control to resolve conflict.

The reactive control effect in the inducer items observed in the participants with PD parallels results by Ruitenberg et al. (20) who found no impairment in reactive control irrespective of participants being tested ON or OFF their dopaminergic medication. However, based on our analysis we would not exclude the possibility of impaired reactive conflict control adaptation in patients with PD, as the effect was explained by improved facilitation in participants with PD, in contrast to reduced interference in HC participants. Ruitenberg et al. (20) reported the difference in Stroop effects irrespectively of the nature of the cost effect (incongruent/congruent items). It would be interesting to learn to what degree the effects in their task depend on improved conflict processing. Future research is needed, to replicate ISPCE manipulation effects in PD in different tasks.

### Interpretation of Electrophysiological Adaptive Control Effects

We found midline-frontal theta modulation to be associated with successful conflict adaptation regardless of the type of control adaptation (both proactive and reactive) in the HC group. We observed a reduction in midline-frontal theta for proactive control adaptation in both the PD and HC group. This is in line with previous studies of proactive control on the CSE (Pastötter et al.(29) and the LWPCE (Chinn et al. (28) that also report reduced conflict theta during conflict processing. However, the group with PD showed evidence of modulation of midline-frontal theta despite their inability to form general context-control associations. Therefore, the presence of conflict theta modulations during conflict processing alone may not suffice to establish intact proactive control adaptation.

We observed a similar modulation of conflict midline-frontal theta in the reactive control adaptation proportion of the task for HC participants. We did not find evidence of conflict theta modulation in participants with PD. In conjunction with their comparatively weaker ability to improve on conflict trials, this suggests that reduced conflict theta in high conflict trials may be a necessary condition for effective reactive control. Our results show the opposite pattern of theta modulation described by Jiang et al. (30). However, the theta cluster Jiang et al. (30) identified was located over midline-posterior electrodes, not midline-frontal electrodes. Moreover, the study was performed in young and healthy participants. Thus, it is conceivable that aging effects may also affect the patterns of control related theta identified in both studies.

In conjunction the results of both experimental manipulations suggest that successful adjustment of control to context, irrespective of reactive or proactive, is associated with a reduction in midline-frontal theta in response to conflicting stimuli. This reduction in theta could reflect more efficient use of conflict resolution resources in control adaptation. When the context indicates a high probability of conflict, less control resources are required to resolve encountered conflict. Vice-versa when conflict expectation is low more control resources are required to resolve encountered conflict.

### Explanations for Deficits Observed in Parkinson’s Disease

Two possible interpretations of the deficit observed in Parkinson’s disease come to mind. The first is the dopamine-overdose hypothesis (51–53). PD is associated with a loss in midbrain dopamine cells. Due to differential neurodegenerative progression in dorsal striatal circuits and ventral striatal circuits (54), dopamine replacement therapy such as Levodopa (L-Dopa) can have divergent effects on cognition. Cognitive functioning reliant on impaired dorsal striatal circuits may profit from dopaminergic medications, whereas functions relying on relatively intact ventral striatal circuitry may display impaired functioning. For example, L-Dopa associated increased tonic and phasic dopamine leads to over-activity of the direct pathway and suppression of indirect pathway, due to differential binding to D1(direct) and D2(indirect) receptors and modifies reinforcement learning behavior. Medicated participants are more inclined to seek rewards, instead of learning from failures i.e., avoiding punishment (55). Similarly, it has been proposed that brain regions associated with conflict monitoring rely more on the spared ventral striatal circuits (56). If proactive control adaptation depends on the ventral striatal circuitry it would be reasonable to expect impaired performance in medicated participants with PD as tested here. To confirm this prediction, future research should assess if performance in unmedicated patients with PD is not impaired in a diagnostic test set.

Another interpretation could be that participants can learn simple stimulus-driven associations but are unable to perform sustained top-down control, informed by learned priors, independent of dopaminergic medication. This is in line with work by Perugini et al. (57), who observed that participants with PD were unable to incorporate prior information during perceptual decision-making, when performing a glass pattern task (similar to the random-dot motion task). Participants learned that a particular response (left or right) was more likely within a given temporal context or item feature and were presented with visual evidence where the degree of visual stochastic noise could introduce conflicting interpretations. Having learned the prior probability of a response option in both task versions, participants with PD were unable to use this information under conditions with high uncertainty, where it would be most relevant. They continued to respond stimulus-driven even under conditions where physical evidence was minimally informative and maximally uncertain. The authors suggest that this reflects the inability of patients with PD to incorporate prior information in their decision-making. However, this could also be interpreted as a failure to engage in control adaptation and continued reliance on uninformed stimulus-driven responding. The failure to use prior information is believed to reflect basal ganglia impairment (58,59), as it was observed irrespective of dopaminergic status (ON/OFF) (58). Moreover, participants with dopamine unresponsive focal dystonia, a disorder with impaired basal ganglia function but unaffected frontal dopamine circuits (58), exhibited similar impairments as participants with PD. The “monochromatic” task (temporal context predicts response) used (57) shares features of proactive control adaptation, whereas the “dichromatic” task (item feature color predicts response) resembles reactive control adaptations. The tasks differ that in the Perugini et al. studies (57,58) participants could predict a concrete response within a given temporal window or feature, whereas in our task they were able to predict conflict. Nonetheless, it is striking how in both, conflict adaptation and informed perceptual decision-making, participants with PD appear to struggle to translate learned probabilistic stimulus information into concrete behavioral adaptations.

### Strength and Limitations

To our knowledge, the present study is the first to evaluate both proactive control and reactive control in PD while controlling for S-R learning. We improved on previous work by having a relatively large sample size (30 participants per group), using state of the art statistical models to assess effects on reaction time distributions, and analyzing EEG frequency correlates of adaptive control in PD. Nonetheless, results need to be viewed in light of some limitations. Our conclusions with regard to reactive control are limited as our diagnostic manipulation was unsuccessful. The feature of the items that we manipulated may have been too abstract. In the future we suggest the manipulation of a more salient feature. Nonetheless, it is unlikely that results in an unbiased diagnostic item set would show improved performance on conflict trials in the participants with PD. Moreover, a failure to identify an ISPCE in the diagnostic or transfer items is not uncommon. For example, Bejjani et al. (60) were unable to find an ISPCE in the diagnostic items. Further, our conclusions of the EEG analysis are limited due to our choice of a short fixation interval. This was done in order to maximize the amount of trials/power to detect an effect. Using the whole trial duration as a baseline allowed us to identify differences in transient modulations of midline-frontal theta, but not sustained changes. Lastly, while our study provides evidence of the state of adaptive control in participants with PD on their dopaminergic medication, it is yet to be conclusively determined to what degree adaptive control abilities are impacted by disease and medication (20,56).

## Conclusion

We demonstrated distinct impairments of proactive-reactive control in participants with PD, when tested on their usual medication. Participants with PD appear to be capable to adjust cognitive control to items directly associated with specific cognitive control demands, but are incapable to form general proactive context-control associations. These adaptations cannot clearly be reinterpreted in terms of intact reactive control or caused by S-R learning. Participants with PD, in contrast to HC participants, failed to regulate cognitive control in the reactive control task in items with specific cognitive control demands. A distinguishing feature may have been the salience in conflict signaling the need of control adaptation. Moreover, results of our elderly control sample highlight that successful reactive and proactive control adaptations, may be accompanied by reduced conflict related midline-frontal theta activity - a common correlate for cognitive control. Unfortunately, the trial number of our EEG data was not large enough to isolate and compare signatures of control adaptations in diagnostic and inducer items separately. In future studies, it would be interesting to identify to what degree midline-frontal theta generalizes to unbiased context-control associations and how it is impacted by age. More studies are required to further dissociate to what degree adaptive control deficits can be explained in terms of PD and medication.

## Data Availability

Due to privacy concerns the data cannot be made publicly available. The experiment, and code used for the analysis and experiment can be found at: https://osf.io/bsn8v/

## Acknowledgments

We would like to thank Veronika Weyer-Elberich for her consultation and helpful suggestions for the statistical analysis and Wolf Pink for his support during the data acquisition. Moreover, we would like to thank Martina Bantel for her helpful feedback during the conception of the task used in this study.

## Author Contributions

Conceptualization: J. Kricheldorff, K. Witt, J. Ficke

Methodology: J. Kricheldorff, K. Witt, J. Ficke, S. Debener

Software: J. Kricheldorff

Validation: J. Kricheldorff

Formal Analysis: J. Kricheldorff

Investigation: J.Ficke, J. Kricheldorff

Resources: K. Witt

Data Curation: J.Ficke, J. Kricheldorff

Writing – Original Draft: J. Kricheldorff, K. Witt

Writing – Review and Editing: J. Kricheldorff, K. Witt, J. Ficke, S. Debener

Visualization: J. Kricheldorff

Supervision: K. Witt

Project Administration: J. Kricheldorff, K. Witt

Funding Acquisition: K. Witt

