## Supplement for "Impaired Proactive Cognitive Control in Parkinson’s disease"

### Supplementary Material – Impaired proactive cognitive control in Parkinson's disease

#### 1. Contrast Matrix

| | $C_{CongruencyCO}$ | $C_{ProportionCO}$ | $C_{InteractionCO}$ | $C_{CongruencyPD}$ | $C_{ProportionPD}$ | $C_{InteractionPD}$ |
|---|---|---|---|---|---|---|
| Incongruent_MI_CO | -0.5 | -0.5 | 0.5 | 0 | 0 | 0 |
| Incongruent_MC_CO | -0.5 | 0.5 | -0.5 | 0 | 0 | 0 |
| Congruent_MC_CO | 0.5 | 0.5 | 0.5 | 0 | 0 | 0 |
| Congruent_MI_CO | 0.5 | -0.5 | -0.5 | 0 | 0 | 0 |
| Incongruent_MI_PD | 0 | 0 | 0 | -0.5 | -0.5 | 0.5 |
| Incongruent_MC_PD | 0 | 0 | 0 | -0.5 | 0.5 | -0.5 |
| Congruent_MC_PD | 0 | 0 | 0 | 0.5 | 0.5 | 0.5 |
| Congruent_MI_PD | 0 | 0 | 0 | 0.5 | -0.5 | -0.5 |

#### 2. Shifted Log-Normal Regression Model and Priors

$$\log(RT_{nm} - \exp(ndt)) \sim Normal(\mu_{nm}, \sigma)$$

$$\mu_{nm} = \alpha + u_{subj[n],1} + u_{Item[m],2} + \beta_1 C_{Congruency_{PD}} + \beta_2 C_{Proportion_{PD}} + \beta_3 C_{Interaction_{PD}} + \beta_4 C_{Congruency_{CO}} + \beta_5 C_{Proportion_{CO}} + \beta_6 C_{Interaction_{CO}}$$

$$ndt = \beta_7 PDGroup + \beta_8 COGroup$$

$$\sigma \sim Normal_+(0, 0.5)$$

$$\alpha \sim Normal_+(6.5, 0.5)$$

$$\beta_1 \sim Normal(0, 0.3)$$

$$\beta_2 \sim Normal(0, 0.3)$$

$$\beta_3 \sim Normal(0, 0.3)$$

$$\beta_4 \sim Normal(0, 0.3)$$

$$\beta_5 \sim Normal(0, 0.3)$$

$$\beta_6 \sim Normal(0, 0.3)$$

$$u_1 \sim Normal(0, 0.3)$$

$$u_2 \sim Normal(0, 0.3)$$

$$\beta_7 \sim Normal(5.3, 0.5)$$

$$\beta_8 \sim Normal(5.3, 0.5)$$

#### 3. Logistic Regression Model and Priors

$$Error_{nm} \sim Bernoulli(\theta_{nm})$$

$$\eta_{nm} = \left(\frac{\exp(\theta_{nm})}{1 + \exp(\theta_{nm})}\right)$$

$$\eta_{nm} = \alpha + u_{subj[n],1} + u_{Item[m],2} + \beta_1 C_{Congruency_{PD}} + \beta_2 C_{Proportion_{PD}} + \beta_3 C_{Interaction_{PD}} + \beta_4 C_{Congruency_{CO}} + \beta_5 C_{Proportion_{CO}} + \beta_6 C_{Interaction_{CO}}$$

$$\alpha \sim Normal(-1.3, 1.5)$$

$$\beta_1 \sim Normal(0, 1.5)$$

$$\beta_2 \sim Normal(0, 1.5)$$

$$\beta_3 \sim Normal(0, 1.5)$$

$$\beta_4 \sim Normal(0, 1.5)$$

$$\beta_5 \sim Normal(0, 1.5)$$

$$\beta_6 \sim Normal(0, 1.5)$$

$$u_1 \sim Normal(0, 1.5)$$

$$u_2 \sim Normal(0, 1.5)$$

#### 4. Results of the logistic regression analysis

| **Parameter** | **PD** | | **HC** | |
|---|---|---|---|---|
| | *M(CI) in %* | *$BIF_{10}$* | *M(CI) in %* | *$BIF_{10}$* |
| | | **LWPCE – Inducer Items** | | |
| Congruency | 2.0(1.2/3.2) | >1000 | 2.6(1.6/4.1) | >1000 |
| Block PC | 0.7(0.2/1.4) | 1.04 | 1.4(0.7/2.4) | 0.75 |
| Interaction | -1.2(-2.4/-0.2) | 2.10 | -3.1(-5.2/-1.6) | 5.88 |

| | | | | |
|---|---|---|---|---|
| **LWPCE – Diagnostic Items** | | | | |
| Congruency | 3.0(1.9/4.5) | >1000 | 2.6(1.6/4.0) | >1000 |
| Block PC | 0.6(0.0/1.4) | 4.36 | 0.6(-0.1/1.4) | 0.71 |
| Interaction | -0.7(-2.1/0.6) | 1.74 | -2.0(-3.7/-0.6) | 2.95 |
| **ISPCE – Inducer Items** | | | | |
| Congruency | 1.1(0.6/2.0) | >1000 | 2.2(1.1/4.0) | >1000 |
| Item PC | 0.5(0.2/1.1) | <0.001 | 0.4(-0.3/1.3) | <0.001 |
| Interaction | -1.7(-3.1/-0.8) | 2.10 | -1.4(-3.3/-0.1) | 0.001 |
| **ISPCE – Diagnostic Items** | | | | |
| Congruency | 1.5(0.8/2.5) | >1000 | 2.8(1.5/4.7) | >1000 |
| Item PC | 0.1(-0.3/0.6) | <0.001 | 0.5(-0.2/1.4) | <0.001 |
| Interaction | -0.2(-1.1/0.6) | <0.001 | -2.1(-4.3/-0.6) | 0.009 |

*Table 1 Results of the logistic regression analysis. Mean estimates and 95 percent credible intervals are provided in percent. The factor congruency reflects the difference between incongruent and congruent items, the factor Block/Item PC the relative difference between MC and MI blocks/items and the interaction reflects the difference in conflict effects (incongruent - congruent) in the MI blocks/items relative to the MC blocks/items. BIF exceeding 1000 or smaller 0.001 are abbreviated for the purpose of making the table legible.*

#### 5. Posterior Predictive Checks

**Error! Reference source not found.**

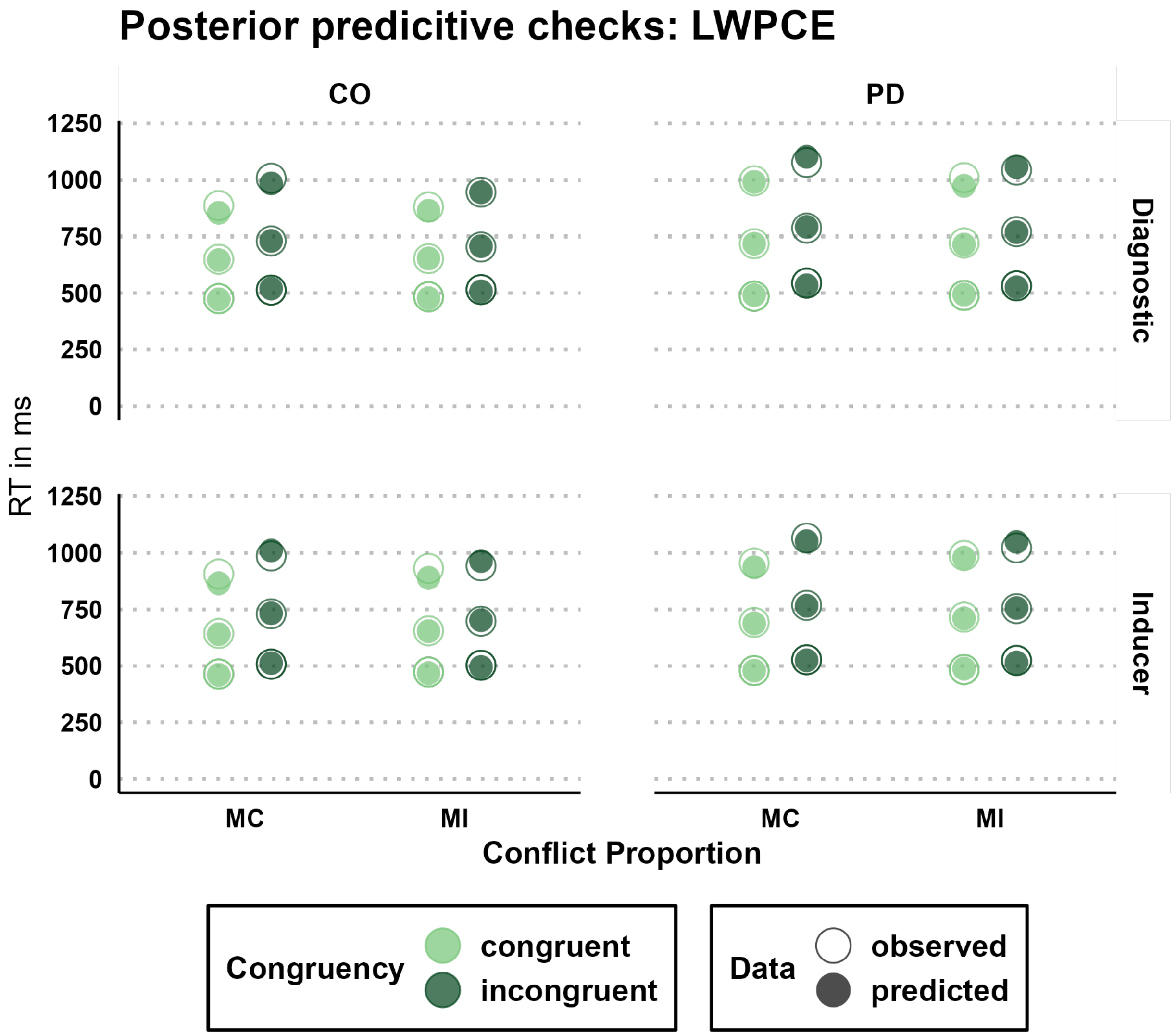


*Figure 1 Posterior predictive checks of reaction times for the analysis of the LWPCE data. Hollow dots depict the observed data, full dots the posterior predictions of the shifted log-normal model. Light green colored dots depict congruent items and dark green dots reaction times to incongruent items. Rows distinguish reaction times to inducer and diagnostic items. Within each panel the upper dot reflects the mean of the upper 90th percentiles of the reaction time distribution, the middle dot reflects the average of the reaction time distribution and the lower dot the mean of the 10th percentile of the reaction time distribution.*

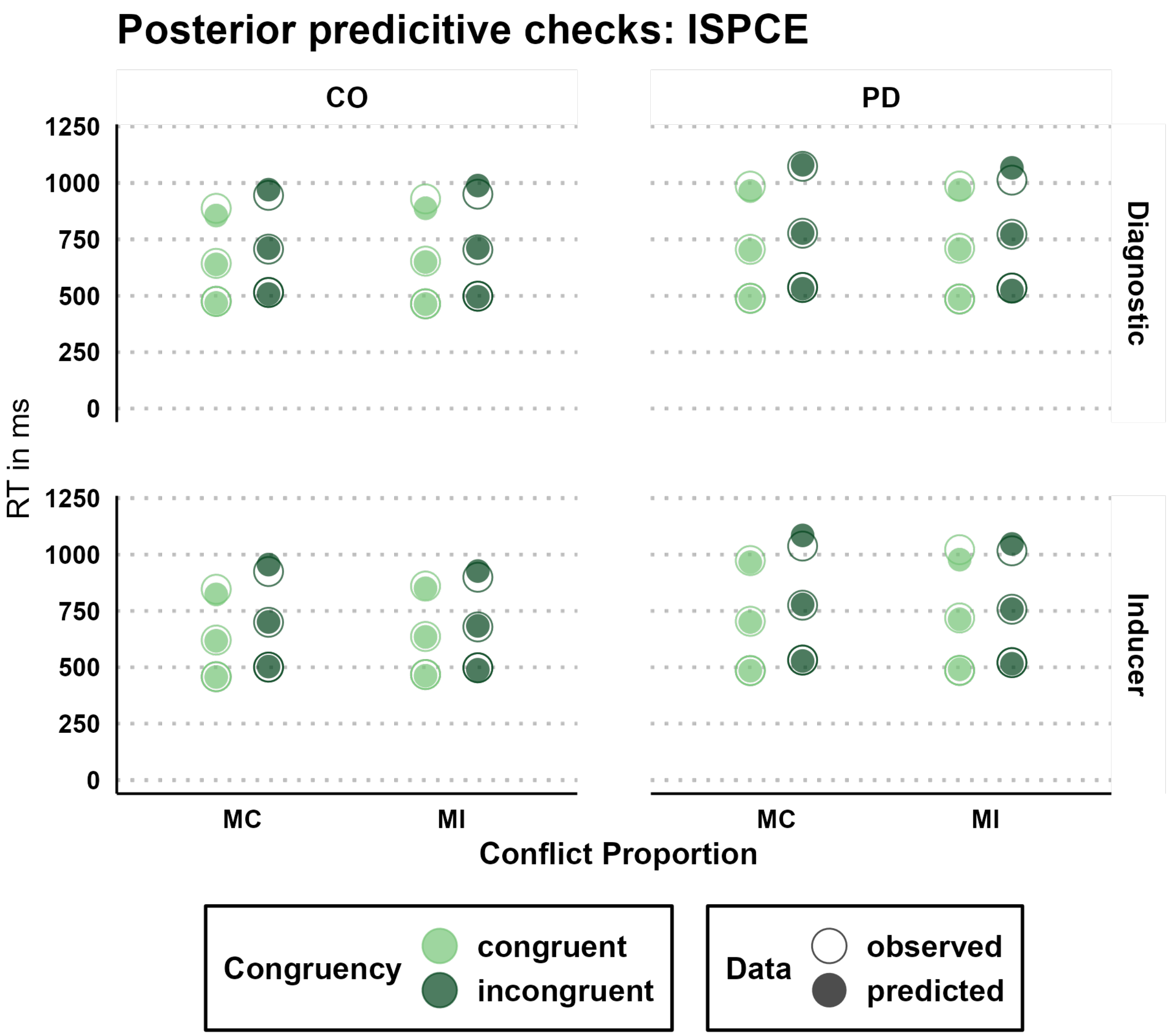


*Figure 2 Posterior predictive checks of reaction times for the analysis of the ISPCE data. Hollow dots depict the observed data, full dots the posterior predictions of the shifted log-normal model. Light green colored dots depict congruent items and dark green dots reaction times to incongruent items. Rows distinguish reaction times to inducer and diagnostic items. Within each panel the upper dot reflects the mean of the upper 90$^{th}$ percentiles of the reaction time distribution, the middle dot reflects the average of the reaction time distribution and the lower dot the mean of the 10$^{th}$ percentile of the reaction time distribution.*

#### 6. Condition Specific Time Frequency Results

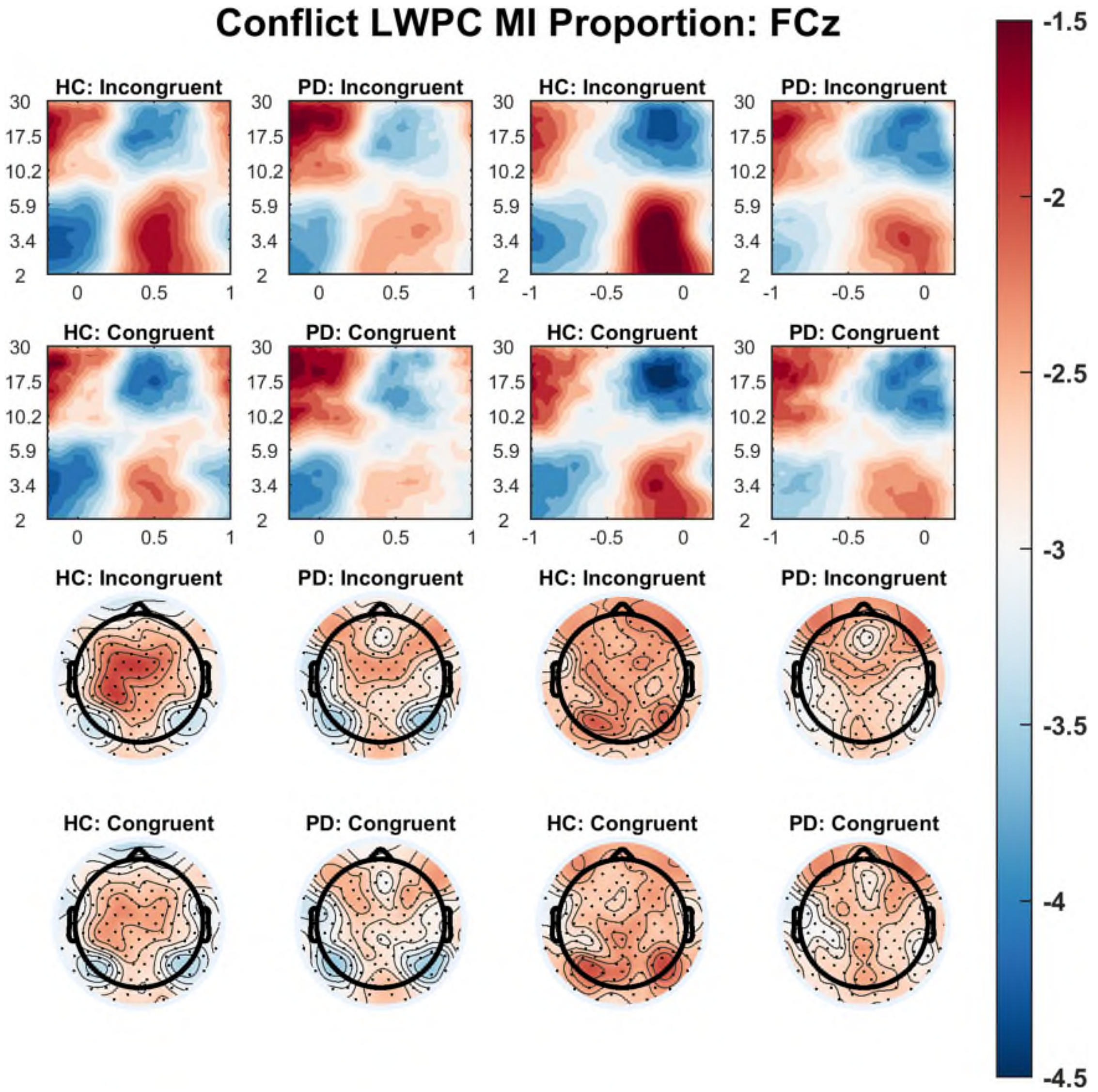


*Figure 3 Estimated marginal mean effects of the time-frequency regression analysis for the LWPCE manipulation of the mainly incongruent block. Data on the two left columns left are plotted in reference to stimulus onset (SL) and data on the two right columns are referenced to the response (RL). The first two rows show the time-frequency results at channel FCz for incongruent and congruent items by group (HC and PD). The last two rows depict the averaged theta power between 0.3 and 0.7s SL and -0.6s to -0.2s RL by congruence and group.*

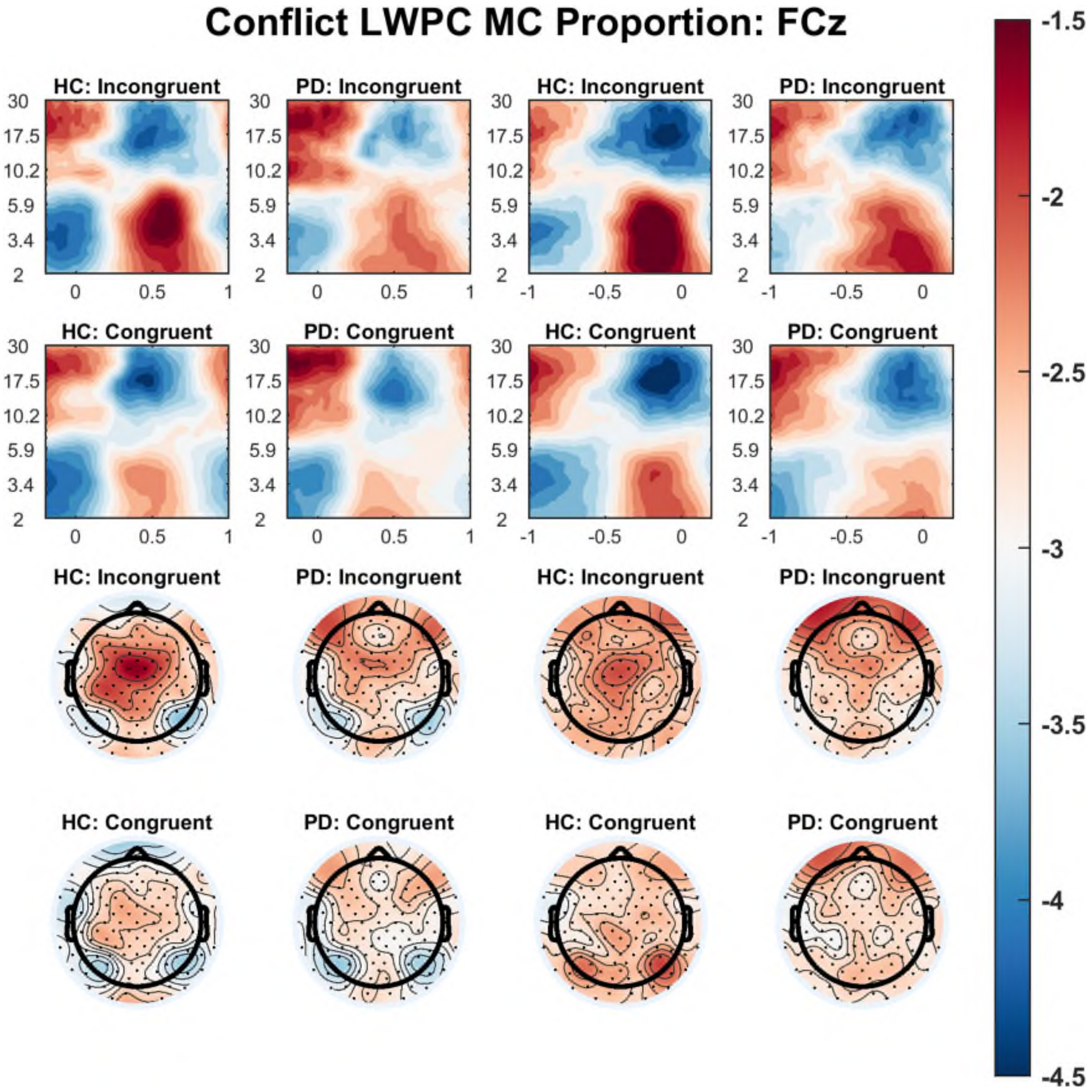


*Figure 4 Estimated marginal mean effects of the time-frequency regression analysis for the LWPCE manipulation of the mainly congruent block. Data on the two left columns left are plotted in reference to stimulus onset (SL) and data on the two right columns are referenced to the response (RL). The first two rows show the time-frequency results at channel FCz for incongruent and congruent items by group (HC and PD). The last two rows depict the averaged theta power between 0.3 and 0.7s SL and -0.6s to -0.2s RL by congruence and group.*

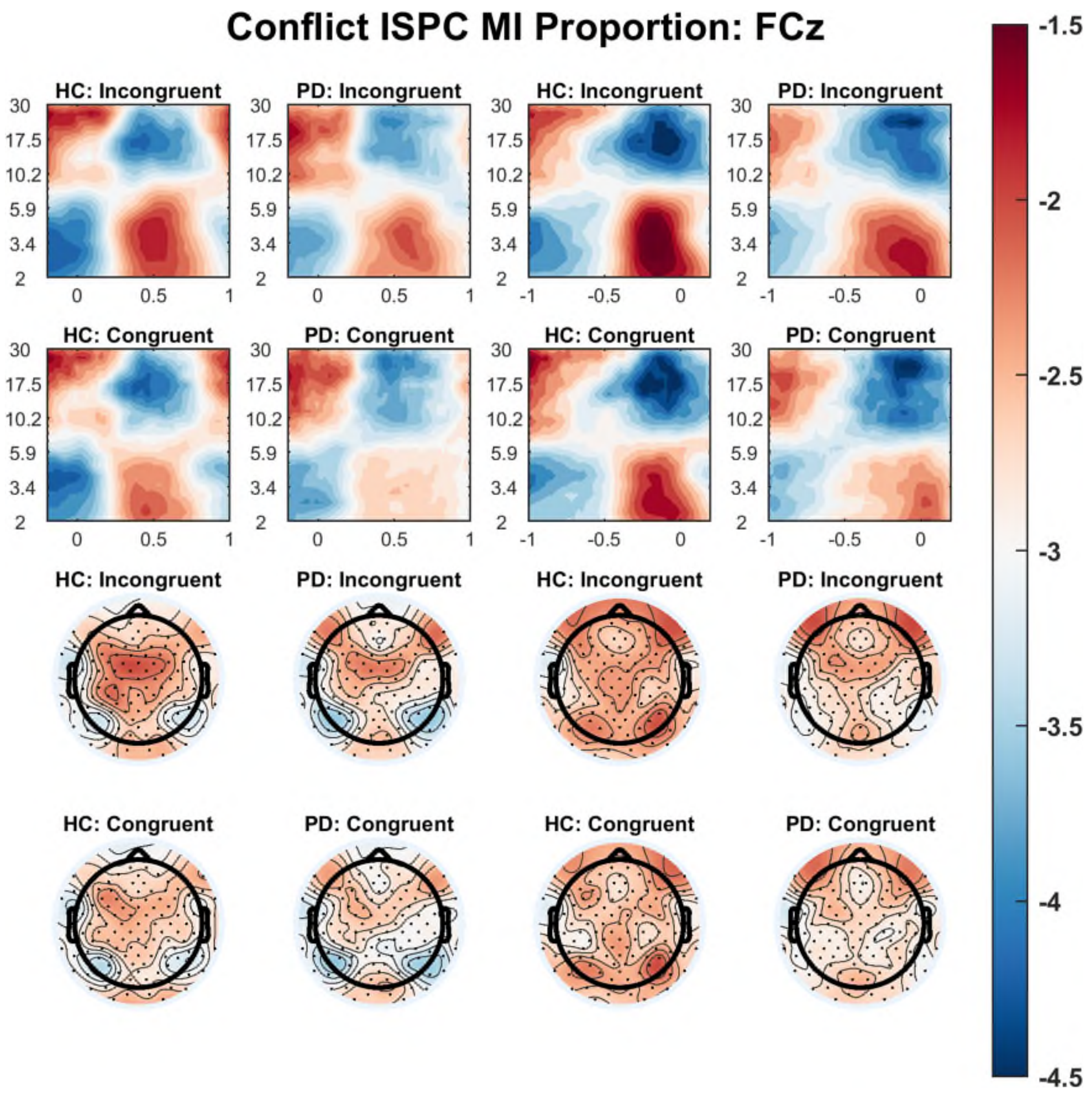


*Figure 5 Estimated marginal mean effects of the time-frequency regression analysis for the ISPCE manipulation of the mainly incongruent items. Data on the two left columns left are plotted in reference to stimulus onset (SL) and data on the two right columns are referenced to the response (RL). The first two rows show the time-frequency results at channel FCz for incongruent and congruent items by group (HC and PD). The last two rows depict the averaged theta power between 0.3 and 0.7s SL and -0.6s to -0.2s RL by congruence and group.*

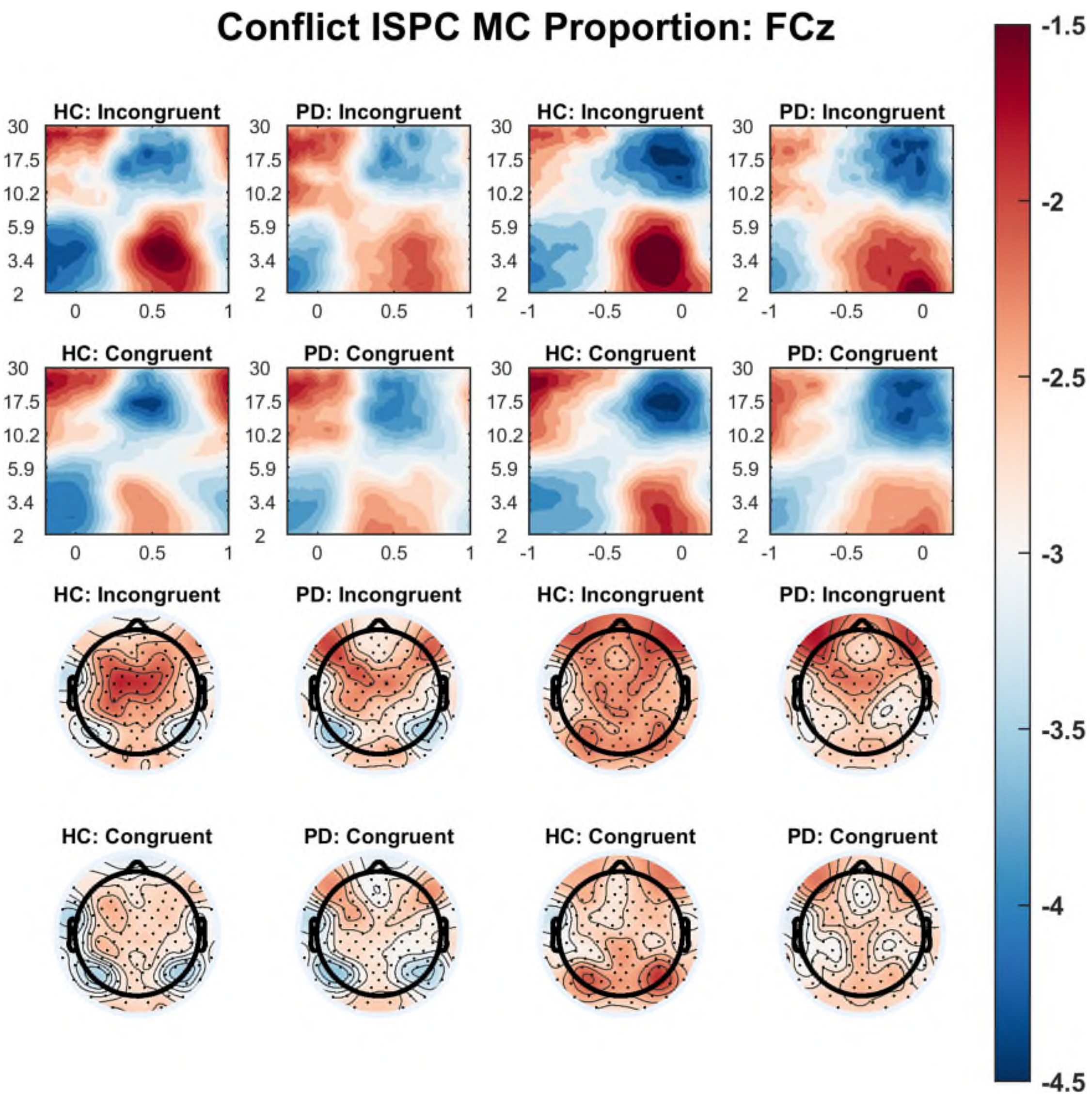


*Figure 6 Estimated marginal mean effects of the time-frequency regression analysis for the ISPCE manipulation of the mainly congruent items. Data on the two left columns left are plotted in reference to stimulus onset (SL) and data on the two right columns are referenced to the response (RL). The first two rows show the time-frequency results at channel FCz for incongruent and congruent items by group (HC and PD). The last two rows depict the averaged theta power between 0.3 and 0.7s SL and -0.6s to -0.2s RL by congruence and group.*
